# Spoken language biomarkers in Turkish-speaking schizophrenia patients: Evidence from linguistic analysis and word embeddings

**DOI:** 10.1101/2025.04.07.25325316

**Authors:** Meltem Çınar Bozdağ, Alper Kumcu, Lütfi Kerem Şenel, Havva Nur Temizkan, Özge Özil, İrem Arslanyürek, Pınar Nazlı Ertekin, Selçuk Candansayar

**Affiliations:** Gazi University, Faculty of Medicine, Department of Psychiatry, Ankara, Türkiye; Ağrı Training and Research Hospital, Department of Psychiatry, Ağrı, Türkiye; Hacettepe University, Faculty of Letters, Department of Translation and Interpreting, Ankara, Türkiye; Ludwig Maximilian University of Munich, Center for Information and Language Processing (CIS), Munich, Germany; Gazi University, Faculty of Medicine, Ankara, Türkiye

**Keywords:** formal thought disorder, language as biomarker, semantic models, speech, word vectors

## Abstract

**Background and Hypothesis:** Schizophrenia (SZ) is considered a “thought disorder”. Therefore, language assessment is crucial in diagnosing SZ. Linguistic analysis and emerging computational language models provide objective biomarkers for diagnosis. Against this background, the main hypothesis is that the language patterns of SZ patients are significantly different from those of healthy controls (HCs) in Turkish, as has previously been shown in other languages.

**Methods:** Speech characteristics of 50 native Turkish-speaking SZ patients were compared with 50 HCs matched for age, sex, length of education, and right/left-handedness. Speech data were collected in 15-minute interviews. The interview recordings were transcribed and analysed for various lexical, syntactic and phonological measures in CLAN and compared for discourse measures using fastText word embedding models.

**Results:** The number of words produced per minute, the number of different words, mean length of utterance, average word frequency, the number of filled pauses, discourse coherence and question-response similarity were lower in the patient group than in the control group. On the other hand, content words/function words ratio, sentence prediction loss, different words/total words ratio, the number of silent pauses, and silent pauses/total speech ratio were higher in the patient group than in the control group.

**Conclusion:** The hypothesis is confirmed. The results from Turkish-speaking SZ patients show similarities with results from other languages from other language families. The findings are important as Turkish is a low-resource and relatively under-researched language in the literature.

The manuscript is under peer review. Please do not cite this preprint.

## 1. Introduction

Certain diagnostic criteria are defined to make a diagnosis in psychiatric practice. The clinician attempts to fit the existing phenomenon into artificially defined diagnostic categories by evaluating the symptoms and signs. In doing so, the clinician examines thought through language, an essential part of the mental status examination. An important part of the assessment of schizophrenia (SZ), which is defined as a “thought disorder”, is the examination of the language used by the individual.

A variety of language abnormalities can be seen in different psychiatric disorders.^1^ However, patients diagnosed with SZ as a formal thought disorder have more frequent and varied language abnormalities compared to patients with other diagnoses. These language abnormalities reflect the underlying thought disorder. Individuals diagnosed with SZ have impairments in semantic, syntactic, and phonological language features.^2–8^ As a result, their ability to understand spoken and written language is impaired, and their quality of life is reduced.^9–11^

Clinicians in psychiatric practice subjectively assess language and thought. Although several scales exist to measure formal thought disorders, objective measures are not yet fully available.^12,13^ Recently, there have been tremendous developments in computational linguistic analysis methods, in line with the developments in machine learning and deep learning. These computational linguistic analysis methods have the potential to be used as biomarkers in psychiatric practice for prediction, diagnosis, treatment selection, prognosis estimation and prevention of recurrent episodes of psychosis.^14–16^ As these methods are developed daily, they can become more precise and accurate in detecting language abnormalities. In the future, they can be used as biomarkers for various disorders.^14,17^ The markers obtained by natural language processing (NLP) methods can be used to scan and classify disorders, predict prognosis, gain insight into the underlying pathophysiology, and develop new treatments.^15^ Studies on this topic are ongoing, and further development of these methods is needed, as the biomarkers defined so far are not accurate and precise enough to be used in routine clinical practice.^15^

Studies utilising linguistic data have largely focused on schizophrenia spectrum disorders (SSDs) because of the abundance of language abnormalities compared to other diagnoses.^17^ Studies have shown a variety of language abnormalities in patients with SSDs, such as abnormalities in semantic memory, intonation, neologisms, difficulties in forming coherent narratives, disruptions in speech cohesion, more frequent referential abnormalities, less syntactic complexity, lower mean length of utterance (MLU), higher type/token ratio (TTR), fewer words, more frequent grammatical errors, more frequent and longer pauses, slower articulation rates, and many others.^4,5,7,8,18–28^ Some of these features have been found to correlate with a variety of clinical features.^7,28–31^ Prediction methods have combined some linguistic features, and promising results have been reported in the literature.^6,7,25^

Most studies in the literature have been conducted with English-speaking patients. Some studies in other languages^6,7,25,28,30,32–36^ need to be extended to define universal and language-specific features. In addition, the samples of most studies were composed of patients with SSDs, including other SZ-like disorders and psychotic disorders. This makes a clinical sample even more heterogeneous, whereas SZ itself is a heterogeneous disorder. Therefore, the present study was conducted in a native Turkish-speaking sample, and the patient group was restricted to those diagnosed with SZ. The main hypothesis is that the speech patterns of patients with SZ will significantly differ from those of HCs in Turkish, as previously shown in other languages. The study’s main aim was to show the differences in daily spoken language data in terms of various language variables (lexical, syntactic, phonological and discoursal) between native Turkish-speaking patients diagnosed with SZ and HC groups.

## 2. Methods

### 2.1. Participants

The sample of the study consists of two groups as SZ patients and HCs. The patient group included native Turkish speakers aged 18-65 years with a diagnosis of SZ according to DSM-5 criteria. Patients were excluded if they had (i) a psychiatric diagnosis other than SZ (including diagnoses other than SZ in SSDs), (ii) a diagnosis of neurological disease that is likely to affect cognitive function, (iii) a history of severe head trauma or neurosurgery, alcohol or drug dependence at the time of assessment or in the past and use of any substance in the past month, and (iv) (uncorrected) hearing or speech impairment. Participants in the control group consisted of healthy individuals with no psychiatric diagnosis at the time of assessment (other than an anxiety or depressive disorder in complete remission, with no symptoms in the past six months and no active use of psychotropic medication). Participants were excluded from the healthy control group if they had at least one first-degree relative diagnosed with a psychotic spectrum disorder. According to the above inclusion and exclusion criteria, 50 patients with SZ were selected from a total of 6,714 patients who presented to the Psychiatric Outpatient Clinic of Gazi University Hospital, Ankara, Türkiye, between 1 October 2022 and 29 August 2023. 50 healthy controls, consisting of individuals who presented for routine check-ups at the Check-up Polyclinic of the same hospital and individuals who volunteered, were matched to the 50 patients regarding age, sex, educational status and right/left-handedness and included in the study. The distribution of sex and handedness was the same between the patient and control groups. The groups had no significant differences in age or length of education. Written informed consent was obtained from all participants. The study was approved by the Ethics Committee for Clinical Research at Gazi University (date: 18/04/2022, approval number: 309). The descriptive characteristics of the sample are shown in Table 1.

**Table 1.** Descriptive characteristics of the patient and the control group

|  | **Patient Group** |  | **Control Group** |  |  |  |
| --- | --- | --- | --- | --- | --- | --- |
| Number of participants | 50 |  | 50 |  |  |  |
| Female/male | 19/31 |  | 19/31 |  |  |  |
| Handedness (right/left) | 45/5 |  | 45/5 |  |  |  |
| Employment (yes/no) | 10/40 | | 46/4 | | $\chi^2 = 52.88$ | $p < .0001$ |
|  | **Mean (SD)** | **Medium (min-max)** | **Mean (SD)** | **Medium (min-max)** | **t** | **p** |
| Age | 46.22 (9.53) | 46.5 (25-64) | 46.52 (9) | 48 (28-65) | 0.16 | .87 |
| Length of education (years) | 10.78 (3.09) | 11 (5-15) | 10.6 (3.23) | 11 (5-15) | 0.28 | .78 |
| Household size in childhood | 5.68 (1.5) | 5 (3-10) | 6.94 (3.53) | 6.5 (2-20) | 2.32 | .02 |
| Age at onset | 25.48 (7.54) | 26 (15-40) |  |  |  |  |
| Duration of illness (months) | 245.04 (93.21) | 240 (60-444) |  |  |  |  |
| PANSS-total | 63.2 (15.25) | 59 (45-105) |  |  |  |  |
| PANSS-P | 13.36 (6.16) | 11 (7-36) |  |  |  |  |
| PANSS-N | 21.12 (6) | 20 (13-35) |  |  |  |  |
| PANSS-G | 28.72 (7.14) | 27 (20-48) |  |  |  |  |

### 2.2. Clinical assessment

The SZ patients were assessed by a senior researcher who took a detailed medical history. Patients were also given a background questionnaire and the Positive and Negative Syndrome Scale (PANSS)^37,38^, which showed that the mean age at onset was 25.48 years and the mean duration of illness was 245.04 months. The total PANSS (PANSS-total) score was 63.2, the positive subscale (PANSS-P) score was 13.36, the negative subscale (PANSS-N) score was 21.12, and the general psychopathology subscale (PANSS-G) score was 28.72. Accordingly, patients with PANSS-N scores greater than PANSS-P scores and PANSS-N scores equal to or greater than the median were classified as negative-dominant. On the other hand, participants with PANSS-P scores greater than PANSS-N scores and PANSS-P scores equal to or greater than the median were classified as positive-dominant. Accordingly, 29 patients were classified as negative-dominant and six as positive-dominant. 68% of the patients had been using atypical antipsychotics (monotherapy or combinations), 6% typical antipsychotics, and 26% combinations of typical and atypical antipsychotics. Descriptive clinical data for the patient group are shown in Table 1.

### 2.3. Data collection procedure

Speech data were collected using a semi-structured interview method. All participants were informed that they would not be called by name, that they would not be asked questions to identify themselves, that they could answer questions as they wished, that they could move on to the next question if they did not wish to answer it, and that they could use fabricated information about place and time to mask the true information if they wished. The interview questions were translated from those used in the study conducted by de Boer et al.^7^ and culturally adapted for the Turkish-speaking sample. The interview questions in the aforementioned study^7^, and thus, in the current study, were designed to encourage participants to speak and were emotionally neutral. That is, the interview did not include questions about the patients’ medical background or any personal information (see supplementary material for the interview questions). The recordings were ensured to last at least 15 minutes. The interviewer did not intervene as long as the participant continued to answer the question. If the participant spoke for more than 15 minutes, the recording was continued until the patient had finished. The questions were asked in a random order so that the participants would feel that it was a more natural conversation. When the patient finished answering, the next question was asked. Two psychiatrists conducted the interviews in a quiet room where distracting stimuli were minimised. Speech data were recorded as .wav files for each participant using a TASCAM DR-40X professional portable recorder and an Audix headset microphone to ensure sound quality suitable for data analysis. Interviewer and participant voices were recorded separately.

### 2.4. Preprocessing of speech data

Audio recordings from 100 participants were transferred to the open-source transcription software CLAN^39,40^, transcribed using the CHILDES-CHAT transcription format^41^, and checked for marking and standardisation against formatting errors. Lexical variables were identified as number of different words (i.e., type), number of total words (i.e., token), type/token ratio, open/closed ratio (OCR; i.e., content word/function word ratio), and average word frequency. Syntactic variables were identified as number of utterances and mean length of utterance. Phonological variables were identified as number of words per minute, number of silent pauses, ratio of silent pauses to total speech (i.e., pause percentage), number of filled pauses, and ratio of filled pauses to total speech (i.e., filled pause percentage). Lastly, discourse variables were identified as discourse coherence, question-response similarity, and sentence production loss.

CLAN markers (i.e., &-, [//], etc.) and elements such as fillers and word fragments marked with these markers were removed from the transcripts; all letters were converted to lower case, all punctuation marks and suffixes separated by quotation marks were removed in the pre-processing stage for the analysis of discoursal language variables. A comprehensive list of ineffective words was created by collecting Turkish stop words from different sources for the ratio of open and closed words. After preprocessing, the total number of content and function words (ineffective words) in the participants’ responses was calculated, and the number of content words was divided by the number of function words. The frequency of the words was calculated based on the Turkish version of Wikipedia, and the frequency of the words used by the participants was calculated accordingly. The semantic similarity between consecutive sentences (phrases written on separate lines in the CLAN) uttered by each participant was calculated for discourse coherence. Word embeddings represented each sentence as a vector in a semantic space. In the word representation method, all words in participants’ responses were represented by a 300-dimensional Turkish fastText word vector,^42^ and sentence vectors were created using the fastText library in Python^43^, which computes the average of the L2 norms of the word embeddings. The average cosine similarities between two consecutive sentences were calculated for each participant. The similarity between the questions asked to the participants and each sentence in the participant’s response was measured. The word representation method was again used to measure similarity. The HuggingFace implementation of the multilingual “mT5-large” language model^44^ was used to calculate the prediction difficulty of the sentences produced by the participants in response to the questions (i.e., sentence prediction loss). Each successive sentence produced by a participant in response to a question was masked, and mT5-large attempted to predict the masked sentence from the question and other response sentences (i.e., masked span prediction). This approach can also combine discourse coherence and question-response similarity analyses. Both sentence prediction loss and question-response similarity metrics are averaged across response sentences. Descriptions of the speech data variables are given in Table 2.

**Table 2.** Linguistic variables analysed in the study and their operational definitions

| **Linguistic variable** | **Definition** |
| --- | --- |
| Number of utterances | An utterance is a chain of words between two pauses in speech, usually ending with a predicate. An utterance can range from a phrase to a whole sentence or even a paragraph, depending on the context and style of speech. |
| Words per minute (WPM) | The number of words produced per minute, which indicates the speech speed |
| Number of types | Total number of different words in a speech |
| Number of tokens | Total number of words in a speech |
| Type/token ratio (TTR) | A high number of repetitions in a speech leads to a lower ratio, while a high number of different words leads to a higher ratio. TTR suggests linguistic richness, lexical variety and expressive creativity in a speech. |
| Mean length of utterance (MLU) | Average number of words per utterance |
| Frequency of silent pauses | The total number of silent pauses is considered an indicator of processing speed. It is influenced by language formulation and planning processes. |
| Percentage of silent pauses | The ratio of silent pauses to the whole speech |
| Frequency of filled pauses | A total number of filled pauses with words like “uh” or “umm”, etc. |
| Percentage of filled pauses | The ratio of filled pauses to the whole speech |
| Open/closed ratio (OCR) | The ratio of open class (content) words to closed class (function) words. Content words are words such as nouns, adjectives, adverbs, etc. and newly derived words. Function words, such as pronouns, have a fixed number and do not accept new members. |
| Average word frequency | The frequency of words in a speech is calculated based on a sample corpus. |
| Discourse coherence | The semantic consistency of sentences in a discourse with one another |
| Question-response similarity | Similarity between the doctor’s question and the patient’s answer. Similarity was calculated separately for each sentence of the patient and the metric is averaged across sentences. |
| Sentence prediction loss | It refers to the loss function that measures the difference between the predicted and actual outputs at the sentence level. Each successive sentence is masked one at a time, and the prediction losses of the masked sentences are computed and averaged using a masked language model (mt5-large). |

### 2.5. Data analysis procedure

The above variables were analysed as formal thought disorder (FTD) indicators. Statistical analyses were performed using R^45^ and Python^43^. Descriptive analyses were performed for clinical data (mean age at onset, illness duration and PANSS scores). Welch’s two-sample t-tests were used to assess differences in linguistic and sociodemographic variables between groups. The chi-squared test was used for categorical variables. Univariate simple linear regression models were fitted with all PANSS scores (positive syndrome, negative syndrome, general psychopathology score and total score), age at onset, duration of illness, type of medication used and demographics as independent variables and the above language variables as target variables. Discourse coherence, question-response similarity, and sentence prediction loss were treated under the heading of discourse variables, and an independent samples t-test was performed in Python^43^ to assess the difference between the groups in terms of these variables. An ordinary least squares regression model was fitted, with sociodemographic and clinical data as independent variables and each discoursal language variable as the target variable, and the predictive power of the variables was analysed.

## 3. Results

### 3.1. Linguistic analysis

SZ patients and HCs were compared using Welch’s two-sample t-test on certain lexical, syntactic and phonological metrics. The results are presented in Table 3, and the selected variables are visualised in Figure 1. The results showed that, compared to HCs, SZ patients spoke slower, produced fewer words and fewer different words (as raw numbers), but produced speech with a higher type/token ratio, shorter utterances, more silent pauses (both as raw numbers and as a percentage of total speech), but fewer filled pauses (as raw numbers). Comparisons of open/closed ratio and average word frequency showed that SZ patients produced speech with a higher open/closed ratio but lower average word frequency.

**Table 3.** Differences between the groups in linguistic variables

| Variable | Level | Patient group |  | Control group |  | %95 CI | *t* | *df* | *p* | *d* | sig. |
| --- | --- | --- | --- | --- | --- | --- | --- | --- | --- | --- | --- |
|  |  | Mean (*SD*) | Medium (min-max) | Mean (*SD*) | Medium (min-max) |  |  |  |  |  |  |
| Number of types | Lexical | 592.3 (159.76) | 607.5 (256-952) | 665.66 (131.7) | 680.5 (164-946) | [15.23, 131.49] | 2.51 | 94.56 | 0.014 | 0.5 | * |
| Number of tokens | Lexical | 1121.3(363.46) | 1130 (411-1966) | 1306.98 (298.61) | 1323.5 (283-2078) | [53.6, 317.76] | 2.79 | 94.44 | 0.006 | 0.56 | ** |
| Type/token ratio | Lexical | 0.54 (0.06) | 0.53 (0.44-0.69) | 0.51 (0.04) | 0.52 (0.44-0.6) | [-0.05, -0.01] | -2.67 | 85.69 | 0.009 | -0.53 | ** |
| Average word frequency | Lexical | 0.001 (0.0002) | 0.001 (0.0005-0.002) | 0.0012 (0.0002) | 0.001 (0.0006-0.002) | [0.00006,0.00025] | 3.3 | 96.76 | 0.001 | 0.66 | ** |
| Open/closed ratio | Lexical | 1.93 (0.27) | 1.93 (1.45-2.84) | 1.71 (0.21) | 1.7 (1.35-2.22) | [-0.32, -0.12] | -4.53 | 91.8 | <.001 | -0.91 | *** |
| Number of utterances | Syntactic | 242.04 (63.7) | 233 (122-408) | 251.56 (52.64) | 248 (166-434) | [-13.68, 32.72] | 0.81 | 94.64 | 0.417 | 0.16 | ns |
| Mean length of utterance | Syntactic | 4.59 (1.03) | 4.62 (2.66-7.04) | 5.32 (0.81) | 5.15 (3.69-7.42) | [0.37, 1.1] | 3.97 | 92.87 | <.001 | 0.79 | *** |
| Words per minute | Phonological | 93.86 (18.55) | 94.83 (43.8-132.99) | 108.82 (14.17) | 110.01 (65.51-140.6) | [8.4, 21.52] | 4.53 | 91.66 | <.001 | 0.91 | *** |
| Number of silent pauses | Phonological | 27.02 (16.59) | 26 (2-70) | 14.88 (8.58) | 14 (2-37) | [-17.4, -6.88] | -4.6 | 73.45 | <.001 | -0.92 | *** |
| Silent pause percentage | Phonological | 1.3 (0.76) | 1.1 (0.24-3.97) | 0.63 (0.4) | 0.57 (0.05-1.97) | [-0.91, -0.43] | -5.57 | 74.67 | <.001 | -1.11 | *** |
| Number of filled pauses | Phonological | 75.06 (48.62) | 61.5 (7-211) | 102.62 (52.11) | 94 (24-228) | [7.56, 47.56] | 2.73 | 97.53 | 0.007 | 0.55 | ** |
| Filled pause percentage | Phonological | 3.49 (1.79) | 3.23 (0.8-8.23) | 4.09 (1.84) | 4.14 (1.26-7.8) | [-0.12, 1.33] | 1.67 | 97.9 | 0.099 | 0.33 | ns |
| Discourse coherence | Discourse | 0.5 (0.05) | 0.51 (0.39-0.59) | 0.53 (0.03) | 0.53 (0.48-0.58) | [0.02, 0.05] | 4.45 | 77.34 | <.001 | 0.89 | *** |
| Question-response similarity | Discourse | 0.44 (0.05) | 0.44 (0.32-0.54) | 0.48 (0.04) | 0.48 (0.37-0.56) | [0.02, 0.06] | 4.32 | 95.46 | <.001 | 0.86 | *** |
| Sentence prediction loss | Discourse | 4.55 (0.23) | 4.52 (4.22-5.15) | 4.43 (0.2) | 4.41 (4.08-5.02) | [-0.2, -0.03] | -2.64 | 90.89 | 0.01 | -0.54 | * |

**Figure 1.**
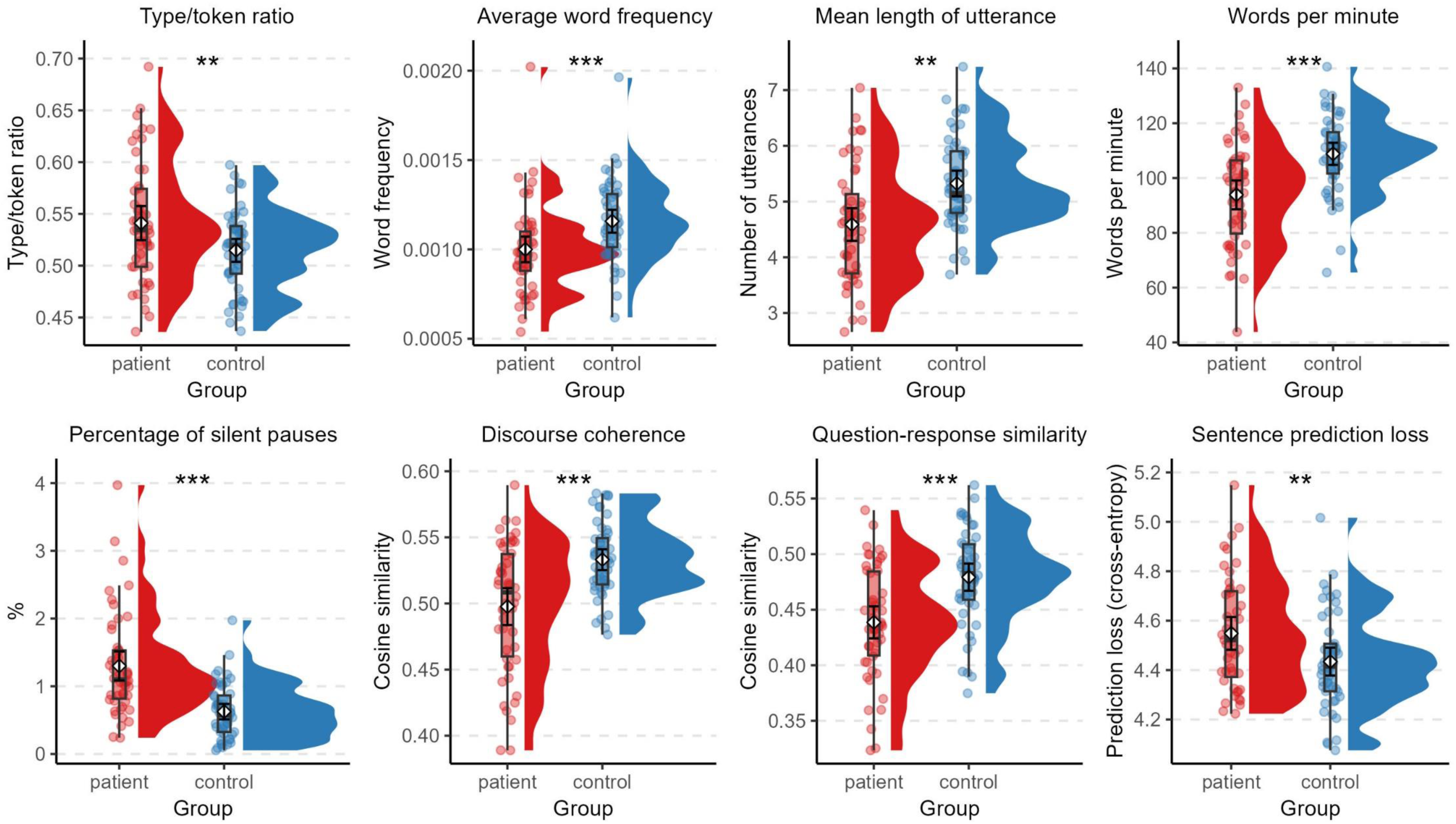
Half violin plots showing differences between the groups in selected linguistic variables

Word embeddings analysis showed that the discourse coherence of the patient group diagnosed with SZ was significantly lower than that of the healthy control group. The similarity between the questions asked to the participants and each sentence of the answer given by the participant was also analysed. Each answer the participant gave was numbered (position 1: 1^st^ sentence of the answer, position 2: 2^nd^ sentence of the answer, etc.), and the degree of semantic similarity between the question and response sentences and between the response sentences themselves was analysed. The variable question-response similarity was found to be significantly lower in the patient group than in the control group on average across positions. The relationship between the distance to the question and the mean similarity score is shown graphically in Figure 2 for the patient and control groups. Analysis with the mT5-large language model showed that the sentence prediction loss was significantly higher in the SZ group than in the control group, indicating that the patients’ responses were less predictable.

**Figure 2.**
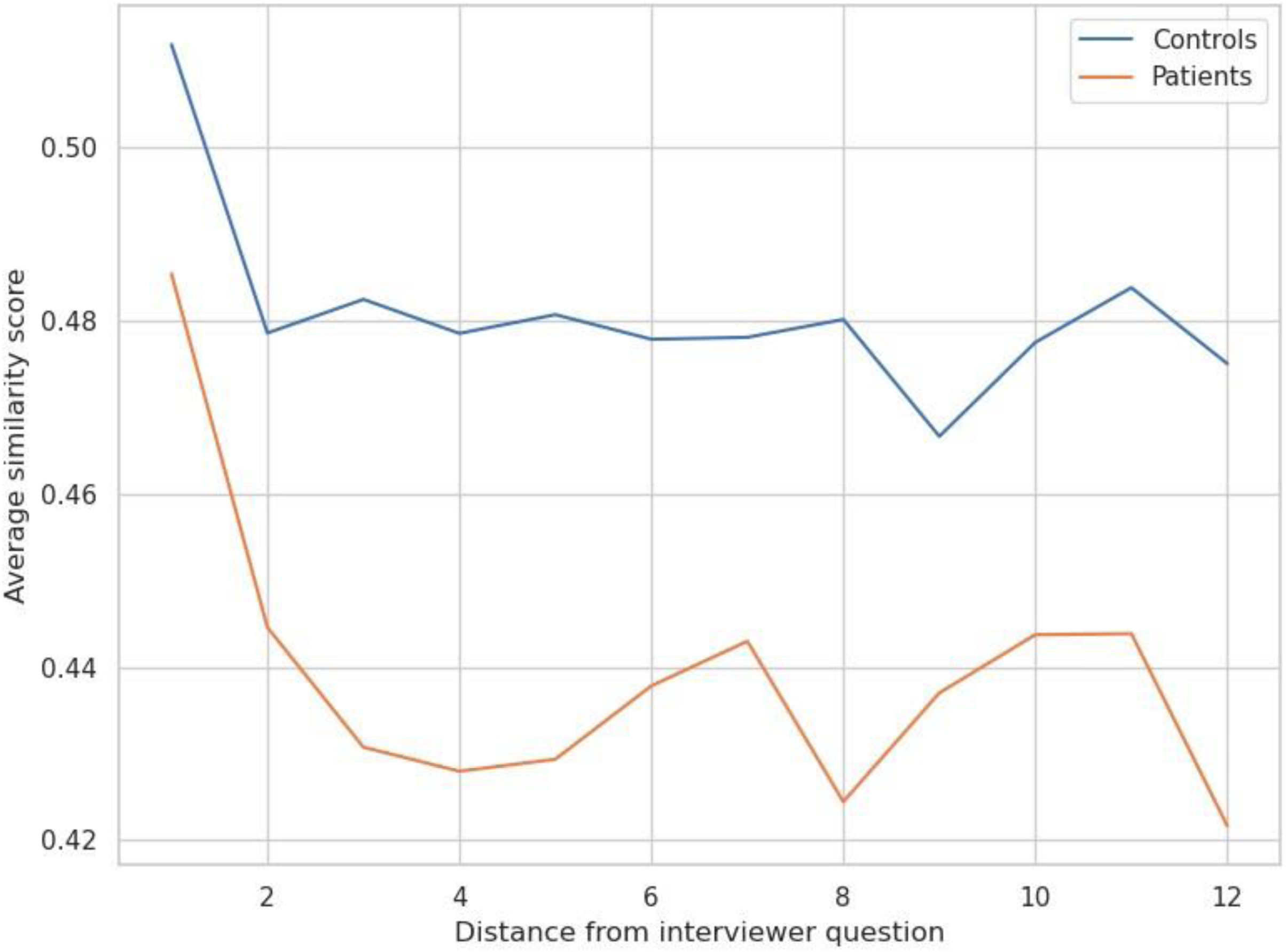
Line chart showing the difference between the groups in question-response similarity across distance to the question

### 3.2. Predictors of linguistic variables

Univariate simple linear regression models were fitted with (1) clinical variables (i.e., all PANNS scores, age of onset, duration of illness, Clozapine use and type of medication) and (2) sociodemographic variables (i.e., age, sex, education, household size in childhood, months without parents) as predictor variables and the language variables analysed above as target variables.

The clinical variable models showed that the PANSS-N scores significantly predicted the number of words produced per minute (WPM), mean length of utterances (MLU), and average word frequency. Patients scoring higher on the PANSS-N produced fewer words per minute, i.e., spoke more slowly; *B* = -0.88, *t* = -2.07, *p* = .04, with shorter utterances; *B* = -0.06, *t* = -2.65, *p* = .01, and rarer words; *B* = -0.00002, *t* = -2.72, *p* = .009. PANSS-N score also significantly predicted discourse coherence and sentence prediction loss. The patients’ responses with higher PANSS-N scores were less coherent, *B* = -0.0036, *t* = -3.38, *p* = .001, and more difficult to predict, *B* = 0.02, *t* = 3.10, *p* = .003. Duration of illness predicted the WPM, discourse coherence, and sentence prediction loss. Accordingly, discourse coherence decreases, and sentence prediction loss increases with the increase in disease duration. Patients who had been ill longer spoke slower; *B* = -0.06, *t* = -2.39, *p* = .02, with lower coherence; *B* = -0.0002, *t* = -2.52, *p* = .02, and their sentences were harder to predict; *B* = 0.0009, *t* = 2.58, *p* = .01.

The models for the sociodemographic variables showed that age, gender, education, and household size in childhood predicted the WPM and the MLU. Age and education also predicted average word frequency and sentence prediction loss. Accordingly, older patients spoke slower; *B* = -0.84, *t* = -3.33, *p* = .002, with shorter utterances; *B* = -0.04, *t* = -3.05, *p* = .004, and rarer words; *B* = -0.00001, *t* = -3.42, *p* = .001, and their sentences were harder to predict; *B* = 0.01, *t* = 2.59, *p* = .01. Female patients spoke slower; *B* = -10.98, *t* = -2.10, *p* = .04, and with shorter utterances; *B* = -0.80, *t* = -2.87, *p* = .006 as compared to male patients. Patients with more years of education spoke faster; *B* = 1.86, *t* = 2.62, *p* = .03, with more frequent words; *B* = 0.00003, *t* = 2.31, *p* = .03, and their sentences were easier to predict; *B* = -0.02, *t* = -2.19, *p* = .03. Finally, household size in childhood positively predicted the number of utterances produced. As the household size increased, so did the number of utterances; *B* = 11.74, *t* = 2, *p* = .05. Selected relationships are shown in Figure 3. None of the other clinical or sociodemographic variables predicted the linguistic variables.

**Figure 3.**
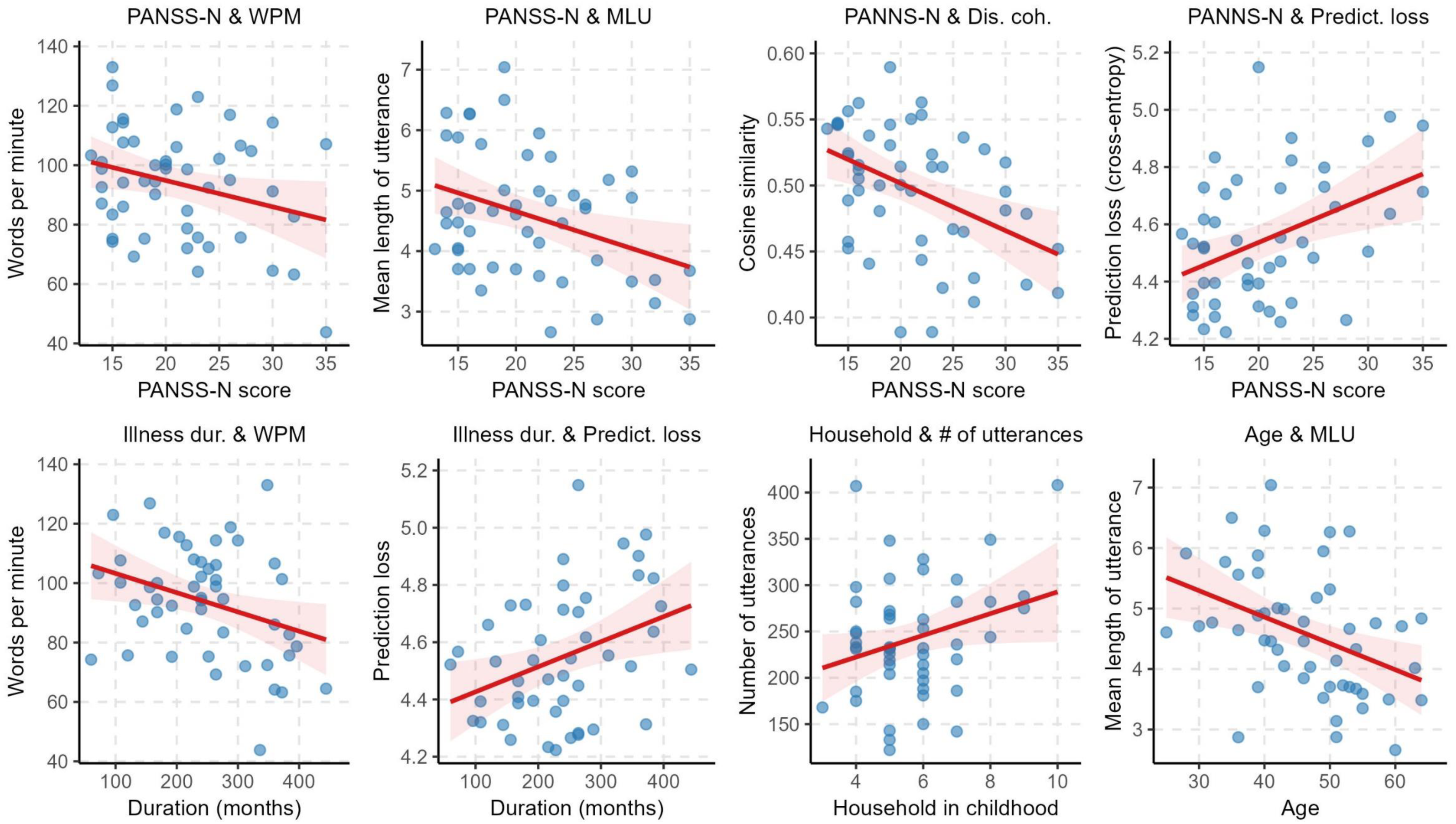
Scatter plots showing the correlations between selected clinical and sociodemographic variables and linguistic variables

## 4. Discussion

The present study aimed to determine whether there are differences in language variables between a group of patients with SZ and a group of HCs. Per this aim, the participants’ spoken language data were analysed for various lexical, syntactic, phonological and discoursal language variables. The SZ and HC groups were matched for age, gender, education and right/left-handedness to minimise confounding factors. The patient group consisted mainly of people who had been ill for more than 20 years and had predominantly negative symptoms. Overall, patients with SZ showed significant differences in a number of language variables compared to the HC group regarding language level.

At the lexical level, the SZ group produced fewer words and fewer different words than the HC group but produced speech with a higher TTR. Although the TTR results may seem counterintuitive, the findings are consistent with the literature.^7,25,28^ It is hypothesised that patients with SZ produce more “unusual” words, possibly due to a disrupted inhibitory mechanism of context-irrelevant linguistic activation, resulting in impaired planning and execution of context-related discourse.^46^ Our study showed that the average word frequency was significantly lower in the SZ group. This may be related to the patients’ lower use of common words and/or a higher use of rare words. Rare words and neologisms are known to be produced by SZ patients in clinical practice. In addition, studies showing lower average word frequency also suggest semantic association and categorisation problems in SZ patients.^47^ Average word frequency should be used with caution as a possible biomarker for SZ, as the literature is mixed.^8,48,49^ OCR was significantly higher in the patient group. As open-class words can accept newly produced words compared to close-class words, OCR is expected to be consistent with a higher TTR in the patient group. It may also be related to patients’ lower use of function words. Conflicting OCR results have been reported previously, with studies finding no significant difference between groups.^7,28^ Further studies and more refined analyses are needed to explain this variable.

At the syntactic level, the SZ group produced speech with a shorter MLU than the HCs. MLU is one of the leading indicators of reduced syntactic complexity. Thus, it can be considered an objective measure of poverty of speech/thought, together with WPM as a function of speaking rate, TTR and pause percentage. The SZ group consists of patients with a long duration of illness and predominantly negative symptoms. Therefore, the shorter MLU in patients aligns with the literature and represents a poverty of speech/thought.^7,28,35,50,51^ At the phonological level, the abovementioned inhibitory dysfunctions are hypothesised to slow down the speech process, resulting in slower production,^46,52^ especially compared to HCs.^7,27,35^ Consistent with this, SZ patients spoke slower than HCs in the present study. Further, the SZ group had significantly more silent pauses, extending the literature.^26,28,53–55^ It has been shown that there are differences in the pattern of pauses in SSDs and that the duration of pauses may be related to negative symptoms.^26,27,31^ Our findings support previous studies because most of our patients had predominantly negative symptomatology. For example, in a neuroimaging study, brain areas associated with speech planning and monitoring showed reduced activation during pauses.^56^ It is thought that these patients have difficulty organising their thoughts and planning speech, resulting in longer pauses.^55,57^ On the other hand, a filled pause is thought to be related to potential error detection during the pre-articulation phase or listener-oriented features of speech.^58–60^ Previous studies have shown fewer filled pauses in patients with SZ,^56,61^ but one study showed no significant difference between groups.^55^ Patients had significantly fewer filled pauses than HCs, but the percentage of filled pauses was not significantly different between groups. Further research is needed to clarify these conflicting results.

At the discourse level, the discourse coherence was significantly lower in the SZ group than in the HC group. This is consistent with both clinical observations and previous studies.^6,20,25,29,36,62,63^ Although a recent study conducted on Turkish found higher semantic similarity in SSD patients, the analysis method used in that study differed from the word embedding method used in the present study.^51^ Discourse coherence is one of the most consistent language variables. Studies have repeatedly shown lower discourse coherence in SZ patients and people at higher risk of psychosis.^64–66^ Therefore, it seems to be the most promising variable for use as a biomarker for patient screening, diagnosis and monitoring. The present study also analysed the similarity between the questions asked of the participants and each sentence of the response given by the participant. The question-response similarity was significantly lower in the patient group than in the HC group. The mean semantic difference between question and response was more pronounced at the beginning of the answers. Unlike our study, there is evidence for lower question-response similarity in SZ patients but gradually increasing differences with each successive sentence.^67^ The sentence prediction loss score was significantly higher in the SZ group than in the HC group, which means that the masked span prediction model produced more errors in predicting masked sentences in the patients’ speech. Overall, findings show less semantic coherence in the speech of the SZ patients.

Lastly, regression models were fitted to show the predictive relationships between sociodemographic, clinical and language variables. Sociodemographic models showed that with increasing age, the speech rate slows down, utterance length shortens, average word frequency decreases, and sentence prediction loss increases. Studies have shown reduced semantic coherence in older SZ patients.^48^ However, older age may also be associated with longer illness duration. Female patients spoke more slowly and with shorter utterances than male patients. One study found lower syntactic language scores in female patients with SZ.^30^ However, as the clinical course and characteristics of female and male patients differ, the available data are insufficient for further interpretation. With increasing years of education, speech rate and average word frequency increase while sentence prediction loss decreases. There is evidence that poverty of thought is more evident among less educated patients.^68^ The clinical models showed that patients with higher scores on the negative syndrome scale spoke more slowly and with shorter utterances. Also, as the PANSS-N score increases, average word frequency and discourse coherence decrease, and sentence prediction loss increases. As illness duration increases, speech rate and discourse coherence decrease and sentence prediction loss increases. These findings are consistent with the literature and clinical evidence.^7,29–31^ Accordingly, the PANSS-N score was found to be an important predictor, possibly because most patients in this patient group had a longer duration of illness and predominantly negative symptoms.

Language deficits in this study and others likely connect to the Theory of Mind deficits and could explain the aetiopathogenesis of psychosis.^69^ If language is seen as a tool for interpreting reality, it may be possible to view schizophrenic thinking as a perception/interpretation deficit.^70^ Incorrectly encoded information in semantic memory may distort the evaluation of new information.^46,71^ Syntactic distortion and semantic distortion in thought production may constantly influence each other in ways that are incompatible with reality.^46^ In this case, when the schizophrenic mind attempts to perceive reality, it misinterprets it, and as it misinterprets, its perceptual errors increase, and the relationship to reality may be broken.^46,70^

The study has limitations. Firstly, the sample size of the groups in this study was relatively small. Secondly, cognitive measures that could potentially confound the results (e.g., attention, memory, working memory and IQ scores) were not included. Thirdly, although the participants’ history of alcohol and drug use was questioned in detail and confirmed by the patients’ relatives, urine drug screening was not carried out. Finally, the medications used by the patients and their side effects could not be analysed due to the small sample size. Antipsychotics, in particular, might be a confounding factor based on reports of different effects of typical and atypical antipsychotics on language variables.^28^ It is important to note that Turkish is an agglutinative language, i.e., grammatical functions are marked by adding suffixes to stems. Thus, a single word can convey what would take a whole sentence in a fusional language such as English. This is something to bear in mind when interpreting TTR and MLU results. To be more precise, morphological variants (e.g., *eve* – “to house”, *evler* – “houses”) of a lemma in the present study (e.g., *ev* – “house”) were considered as different words in the TTR analysis. As MLU was calculated on a word basis, a calculation based on morphemes per utterance may give a different picture.

The present study also has notable strengths. To the best of our knowledge, this is one of the first studies to measure language variables in native Turkish-speaking SZ patients. Importantly, unlike most studies, the patient group was restricted to the diagnosis of SZ only, not SSDs. This provides a more homogeneous group of patients and reduces confounding factors. In addition, the study was the first to use sentence prediction loss analysis in the analysis of speech in SZ, contributing to the literature on semantic analysis models.

In conclusion, the present study demonstrates the differences in spoken language between SZ patients and HCs regarding various lexical, syntactic, phonological and discoursal language variables. The SZ group showed significant differences in speech rate, TTR, MLU, variables related to pauses, open/closed ratio, average word frequency, discourse coherence, question-response similarity, and sentence prediction loss compared to HCs. The results from Turkish-speaking SZ patients show similarities with results from other languages from other language families. The language variables analysed in this study have potential as biomarkers for the quantitative measurement of formal thought disorder. Although current studies tend to focus on the accuracy of a model’s predictive performance, there is a potential that language models may help understand the aetiopathogenesis of the disorders. Further studies with large sample sizes are needed with Turkish-speaking participants to determine the universal and language-specific features of the language variables in SZ and to elucidate the sociodemographic characteristics and clinical details of language performance in patients with SZ.

## Data Availability

All data produced are available online at https://osf.io/x7qd2/?view_only=1b746534b8a24a06b9a19935e6ceebd8

## Acknowledgements

The authors would like to acknowledge the students and research assistants responsible for obtaining and transcribing the conversations used in our study, as well as all study participants.

## Supplementary material

Supplementary material is available at https://osf.io/x7qd2/?view_only=1b746534b8a24a06b9a19935e6ceebd8

## Data availability

The authors confirm that all data supporting the findings of this study are available within its supplementary material.

## CRediT authorship contribution statement

**Meltem Çınar Bozdağ:** Conceptualization, Data curation, Investigation, Methodology, Project administration, Writing – original draft, Writing – review and editing. **Alper Kumcu:** Formal analysis, Methodology, Software, Visualization, Writing – original draft, Writing – review and editing. **Lütfi Kerem Şenel:** Formal analysis, Methodology, Software, Visualization, Writing – original draft, Writing – review and editing. **Havva Nur Temizkan:** Data curation, Investigation. **Özge Özil:** Data curation, Investigation. **İrem Arslanyürek:** Data curation, Investigation. **Pınar Nazlı Ertekin:** Data curation, Investigation. **Selçuk Candansayar:** Conceptualization, Supervision, Project administration, Resources, Funding acquisition.

